# Integrating planetary health and environmental justice into high school construction career education: protocol for a randomized controlled trial of the Ecosystem Justice Translator

**DOI:** 10.64898/2026.07.09.26357686

**Authors:** Devan Cantrell Addison-Turner, Gretchen C. Daily

## Abstract

**Introduction:** Climate change disproportionately affects Disadvantaged Communities, yet construction workforce education rarely addresses interconnected pathways linking energy efficiency, nature exposure, and public health. Green-blue infrastructure delivers co-optimized benefits: reducing building energy consumption 15–30% while decreasing heat-related mortality by approximately 3.9% per degree Celsius of urban cooling (Gasparrini et al., 2017)— epidemiological benchmarks that inform the dose-response functions embedded in the Ecosystem Justice Translator (EJT). This protocol describes, to our knowledge, the first randomized controlled trial evaluating a curriculum intervention designed to develop planetary health competencies and environmental justice awareness among high school students pursuing construction careers.

**Methods and analysis:** This two-arm, parallel-group randomized controlled trial targets enrollment of N=200 high school students (ages 14–18) from construction career pathway programs in the San Francisco Bay Area (over-recruitment target N=250; 25% buffer for attrition). Students are individually randomized 1:1 to intervention (Community-Centered Design curriculum integrating the Ecosystem Justice Translator) or control (traditional Virtual Design and Construction curriculum), stratified by school site using block randomization. The 6-month intervention features the Ecosystem Justice Translator (EJT)—a computational system using large language models to translate community health equity concerns into quantifiable investment priorities. The EJT’s 51-theme health equity taxonomy was derived from validated public health frameworks (Centers for Disease Control and Prevention [CDC] Social Vulnerability Index, Environmental Protection Agency [EPA] EJScreen, Healthy People 2030). Primary outcome is Health-Integrated Equity Consciousness Index (HI-ECI), measured at baseline, 3, 6 (primary endpoint), and 12 months. Analysis uses intention-to-treat linear mixed-effects models with random intercepts for participants. The minimum required sample (n=26 per arm; G*Power, two-tailed α=0.05, 80% power, Hedges’ g=0.80) is exceeded by enrolled N=200, which provides >99% power at Hedges’ g=0.80 and supports multi-site confirmatory factor analysis.

**Ethics and dissemination:** This protocol has been approved by Stanford University Institutional Review Board (IRB eProtocol #84369, approved February 13, 2026). Parental consent from a parent or guardian and written assent from each student participant are required prior to enrollment. All instruments, curriculum materials, and EJT source code will be released open-source under CC BY-NC-SA 4.0, permitting free use for educational, research, and non-profit purposes, concurrent with primary publication. Commercial licensing may be pursued separately through Stanford University Office of Technology Licensing (OTL docket S25-565).

**Trial registration:** ClinicalTrials.gov NCT07315919. Pre-results. Protocol version 4.1, July 2026.

**STRENGTHS AND LIMITATIONS OF THIS STUDY:**

- Individual randomization within school sites incorporates three structural contamination mitigations: mandatory active engagement with the intervention software, blinded outcome assessors, and temporal separation where the same instructor teaches both arms.
- A two-phase psychometric validation strategy—pilot exploratory factor analysis followed by confirmatory factor analysis with measurement invariance testing across school sites in the main trial—establishes the primary outcome’s factor structure and reliability.
- The single geographic context (San Francisco Bay Area) limits generalizability of findings to other regions and career technical education program structures.
- The primary and secondary outcome measures (HI-ECI and related indices) are novel and lack established normative benchmarks, so their psychometric properties will only be confirmed during the pilot phase.
- Curriculum delivery by trained career technical education instructors, rather than research team members, introduces fidelity variability that may attenuate observed effects relative to a tightly controlled efficacy trial.

## INTRODUCTION

### Background and rationale

The built environment profoundly shapes human health through interconnected pathways linking energy systems, nature exposure, and physiological outcomes. Buildings account for approximately 40% of global energy consumption and carbon emissions, while urban heat islands—intensified by impervious surfaces and waste heat—elevate ambient temperatures by 3–7°C in disadvantaged neighborhoods compared to affluent areas.^1,2^ This thermal inequity translates directly into health disparities: heat-related mortality, cardiovascular stress, and respiratory illness concentrate in communities with the least access to cooling infrastructure and green space.^3^

Green-blue infrastructure investments—urban tree canopy, green roofs and walls, bioswales, rain gardens, and parks—address this nexus through multiple synergistic pathways. Energy efficiency gains emerge from evapotranspirative cooling, shading, and insulation: meta-analyses demonstrate 15–30% reductions in building cooling energy demand from strategic urban greening.^4,5^ Nature exposure benefits operate through stress reduction via hypothalamic-pituitary-adrenal axis modulation,^6^ physical activity promotion,^7^ immune function enhancement through phytoncide exposure,^8^ and attention restoration:^6^ systematic reviews confirm 0.12 disability-adjusted life years (DALYs) averted per hectare of accessible greenspace within 300 meters of residences, with 28% lower odds of depression.^9,10^ Direct health protection results from urban cooling (3.9% mortality reduction per 1°C temperature decrease), particulate matter removal via dry deposition, and flood injury prevention through stormwater retention.^11,12^

Federal legislation including the Inflation Reduction Act (Pub. L. 117-169) and Bipartisan Infrastructure Law (Pub. L. 117-58) directed hundreds of billions toward climate and clean energy investments with equity mandates for Disadvantaged Communities.^13^ Yet no validated tools exist to help planners, designers, or construction professionals optimize investments across energy efficiency, nature exposure, and health equity dimensions simultaneously. Existing tools either exclude community voice (InVEST), focus on burden rather than investment optimization (CalEnviroScreen), or lack ecosystem service integration (EJScreen).^14,15^ The CDC SVI (version 2022) and EPA EJScreen data were downloaded and archived locally prior to December 2025; the trial’s methodology does not depend on continued active federal program access. Where federal sources are unavailable, California-equivalent fallbacks are pre-specified: CalEnviroScreen 4.0 (California OEHHA) and the California Healthy Places Index (Public Health Alliance of Southern California).

The EJT addresses this gap by operationalizing three dimensions of environmental justice theory simultaneously in a single computational system. Distributive justice (fair allocation of environmental benefits across communities) is operationalized through the Social Vulnerability Index (SVI)-weighted benefit calculation in the Ecosystem Service Health Integration (ESHI) module. Procedural justice (meaningful community participation in investment decisions) is operationalized through the CVET module, which extracts and centers community voice as the highest-weighted input (40%) in the Environmental Justice Investment Prioritization (EJIP) formula. Recognition justice (acknowledgement of historically marginalized knowledge and identity) is operationalized through six dedicated recognition justice themes in the 51-theme taxonomy, ensuring that cultural respect, traditional ecological knowledge, and anti-displacement concerns are structurally represented in every investment priority calculation. This tripartite operationalization draws on Fraser and Honneth’s theory of justice (2003)^16^ and is, to our knowledge, the first computational implementation of all three justice dimensions in an infrastructure investment prioritization system.

The construction industry employs over 7.5 million workers in the United States, with demand projected to grow 4% annually through 2032 (Bureau of Labor Statistics Standard Occupational Classification 47-0000, Construction and Extraction Occupations).^17^ High school career technical education (CTE) programs represent a critical entry point for developing workforce competencies. However, current construction curricula rarely address environmental justice or planetary health concepts, missing opportunities to prepare the next generation to deliver equitable infrastructure. To our knowledge, no validated, equity-integrated educational instrument exists to assess whether CTE construction students develop the infrastructure literacy needed to recognize environmental health inequities, design equity-informed solutions, and advocate for community-centered investment.^18^ While the intersection of environmental justice and the built environment is relevant across multiple CTE clusters—including Agriculture, Food and Natural Resources; Architecture and Construction; and Science, Technology, Engineering and Mathematics—construction career pathways were prioritized for this initial trial for three reasons: (1) construction workers directly implement or foreclose green-blue infrastructure investments, creating the most proximal pathway from workforce competency to community health outcome; (2) construction CTE programs serve a predominantly low-income, male, and minority-identifying student population, aligning with Justice40 equity priorities; and (3) the EJT was developed in partnership with construction CTE programs, ensuring contextual validity of curriculum materials and outcome measures. Extension to adjacent CTE clusters is planned as a subsequent research phase.

This protocol describes the evaluation of the Ecosystem Justice Translator (EJT)—a computational assessment platform that translates community voices into standardized infrastructure literacy metrics—through a randomized controlled trial conducted in anticipated collaboration with Oakland Unified School District (OUSD), the West Oakland Environmental Indicators Project (WOEIP), and the Santa Clara County Construction Careers Association (S4CA), pending formal agreements.

### Theoretical framework

Three frameworks guide this project. Freire’s critical consciousness pedagogy establishes that genuine education requires naming, analyzing, and acting against structural oppression.^19^ Applied to infrastructure, students must learn not only how to build but who benefits from what gets built and how design decisions perpetuate or disrupt health inequities. Mezirow’s transformative learning theory provides the mechanism: disorienting dilemmas—in this context, community health data revealing that students’ own neighborhoods bear disproportionate infrastructure burdens—trigger critical reflection producing perspective transformation and behavioral change.^20^ Ostrom’s social-ecological systems framework provides the scale: infrastructure literacy must operate across building, neighborhood, and community scales simultaneously, because infrastructure decisions at each scale shape health outcomes at the others.^21^

These frameworks converge in the HI-ECI construct. Problem Recognition operationalizes Freire’s naming^19^—students identify how infrastructure conditions produce health inequities. Design Integration operationalizes Mezirow’s transformation^20^—students apply equity-weighted criteria to technical decisions. Advocacy Activation operationalizes Ostrom’s polycentric action^21^— students develop capacity to communicate community health priorities across governance scales.

#### Theory of change

If CTE students encounter community health data revealing infrastructure inequities in their own neighborhoods (disorienting dilemma), and if curriculum supports critical reflection on how technical decisions perpetuate or disrupt those inequities (conscientization), then domain-specific infrastructure literacy (HI-ECI) will increase measurably relative to students receiving traditional instruction without equity content. This chain generates testable predictions: (1) HI-ECI gains will be largest on Problem Recognition, most directly activated by community data exposure; (2) gains will be sustained at 12-month follow-up because perspective transformation is more durable than surface learning; (3) effects will be largest for students in highest-burden census tracts, where disorienting dilemmas are most proximal.

### Explanation for choice of comparator

The control condition (traditional Virtual Design and Construction [VDC] curriculum) was selected because it represents current standard practice in construction career pathway programs across California and nationally. Traditional VDC curricula cover building codes, construction safety, blueprint reading, computer-aided design (CAD), materials science, and carpentry without explicit health equity, environmental justice, or planetary health content. This comparator allows assessment of the incremental benefit of integrating these concepts beyond existing technical training, providing results directly applicable to program administrators and curriculum developers considering modifications.

### Objectives and hypotheses

#### Primary objective

To determine whether a Community-Centered Design curriculum integrating the Ecosystem Justice Translator improves high school construction students’ health equity consciousness (measured by HI-ECI) compared to traditional VDC curriculum at 6-month post-intervention.

#### Secondary objectives

(1) Assess treatment effects on community health impact recognition (CHIRS) and project health impact assessment (P-HIAS) scores at 6 and 12 months; (2) Evaluate psychometric properties (reliability, validity, factor structure) of the HI-ECI instrument through two-phase validation; (3) Characterize implementation fidelity and participant engagement using process measures; (4) Explore heterogeneity of treatment effects across pre-specified student subgroups defined by baseline environmental burden (CalEnviroScreen 4.0 census tract score).

#### Hypotheses

We hypothesize that: (H1) intervention group participants will demonstrate significantly higher HI-ECI scores compared to control group participants at 6-month post-intervention (primary); (H2) intervention effects will be sustained at 12-month follow-up; (H3) intervention group will show superior CHIRS and P-HIAS scores (secondary); (H4) treatment effects will be larger among students residing in census tracts with higher CalEnviroScreen cumulative burden scores (exploratory).

Hypothesis H4 is the direct empirical test of the third prediction of the theory of change: that disorienting dilemmas are most proximal for students living in highest-burden census tracts, producing the largest perspective transformation and therefore the largest HI-ECI gains. This prediction is theoretically grounded in Mezirow’s account of disorienting dilemmas as experiences that contradict existing frames of reference: students whose own neighborhoods appear in the EJIP investment map as highest-priority for green-blue infrastructure face a more immediate and personally relevant disorienting dilemma than students in lower-burden tracts. The heterogeneous treatment effects analysis using CalEnviroScreen 4.0 quartiles therefore serves a dual function: it is both a pre-specified exploratory analysis and a theory-driven test of the core mechanism of the Freire–Mezirow theoretical framework underpinning the EJT curriculum.

## METHODS AND ANALYSIS

### Trial design

This is a two-arm, parallel-group, superiority randomized controlled trial with 1:1 allocation ratio comparing Community-Centered Design curriculum (intervention) to traditional VDC curriculum (control). Students are individually randomized within participating school sites, stratified by school site using block randomization with randomly varying block sizes (4, 6, 8). The trial follows SPIRIT 2013 guidelines^22^ and incorporates SPIRIT-AI 2020 extension^23^ for artificial intelligence-based interventions and CONSORT-Equity 2017 extension^24^ for health equity reporting. The trial is registered on ClinicalTrials.gov (NCT07315919) prior to enrollment.

**Supplementary materials** Supplementary File 1 provides the completed SPIRIT 2013 and SPIRIT-AI checklists. Supplementary File 2 provides the Technical Supplement describing EJT algorithms, dose-response functions, and scoring procedures. Supplementary File 3 provides the Stanford University Institutional Review Board approval letter (eProtocol #84369). Supplementary File 4 provides IRB-approved parental consent, student assent, and adult consent forms in English and Spanish.

### Plain-language summary

This study tests whether teaching high school construction students about environmental justice and community health—using a computational tool called the Ecosystem Justice Translator— improves their understanding of how buildings affect health equity. Individual students within each participating school are randomly assigned to receive the new curriculum or continue with standard construction training. Student understanding is measured at the start, at 3 months, 6 months, and 12 months using standardized assessments. The study takes place in Oakland, California public high schools serving predominantly low-income students of color in communities disproportionately burdened by environmental hazards.

### Study setting

The trial is conducted in anticipated collaboration with Oakland Unified School District (OUSD), a large urban district serving predominantly low-income students of color across East and West Oakland—communities designated as Disadvantaged Communities (DACs) under California law. Target enrollment spans participating high schools offering construction-focused CTE programs approved under California’s CTE Standards and Framework. Sites were selected to represent demographic diversity, including schools serving DACs. Anticipated collaborations with WOEIP and the Santa Clara County Construction Careers Association (S4CA), pending formal agreements, would provide classroom access, participant recruitment, and community liaison support. Upon agreement, WOEIP would hold decision-making authority over community data interpretation, dissemination framing, and advisory veto rights over findings that conflict with community-defined priorities. Specific school site names will not be disclosed in any publication or presentation without written authorization from OUSD district administration and the relevant school principal. Participating schools are identified only by site number and aggregated demographic characteristics per CONSORT-Equity 2017 reporting guidelines. Confidentiality protections extend to participating institutions: no school will be identified in publications, conference presentations, or public reports without explicit written consent from the institution.

### Eligibility criteria

#### Participant inclusion criteria

(1) Age 14–18 years at enrollment; (2) Current enrollment in a participating construction career pathway program (minimum 2nd semester); (3) Ability to participate in 6-month curriculum during regular CTE class periods; (4) Written informed consent from parent/guardian for participants under 18 years; (5) Written assent from student participant; (6) Ability to complete assessments in English (with accommodations as needed).

#### Participant exclusion criteria

(1) Prior completion of formal environmental justice curriculum within the past 12 months; (2) Expected inability to complete study assessments due to planned relocation or program withdrawal; (3) Concurrent enrollment in another research study involving educational interventions.

### Intervention

The intervention is a 6-month (24-week) Community-Centered Design curriculum integrating the Ecosystem Justice Translator (EJT), delivered during regular CTE class periods (approximately 4 hours weekly, 96 contact hours total). The curriculum follows TIDieR reporting guidelines.^25^

#### EJT system description

The EJT is a web-based computational system comprising four integrated modules:

#### Community Voice Equity Translation (CVET)

Processes qualitative community input using a large language model (the most recent generally available Claude model, Anthropic; version-locked prior to enrollment) adapted to a 51-theme health equity taxonomy derived from validated public health frameworks (CDC SVI, EPA EJScreen, Healthy People 2030) through structured prompt engineering with temperature-scaled softmax calibration (T=0.2; see**Technical Supplement Section 2.1.2**for full fine-tuning specification, system prompt structure, version-locking procedure, and model deprecation contingency plan). The system prompt is version-locked, Secure Hash Algorithm 256-bit (SHA-256) hashed, and archived on the Open Science Framework (OSF) concurrent with trial commencement to enable exact replication.

#### Ecosystem Service Health Integration (ESHI)

Integrates validated InVEST model outputs (Urban Cooling, Stormwater Retention, Nature Access) with peer-reviewed epidemiological dose-response functions (Gasparrini 2017 for heat mortality, Hoek 2013 for PM2.5, Gascon 2016 for mental health). Applies CDC Social Vulnerability Index weighting to prioritize benefits to high-vulnerability populations.

#### Environmental Justice Investment Prioritization (EJIP)

Generates ranked investment priorities using multi-criteria optimization: community voice (40%), health benefits (30%), equity indices (20%), feasibility (10%). Weights are user-adjustable.

#### Uncertainty, Bias, and Risk Quantification (UBR)

Propagates parameter uncertainty via Monte Carlo simulation (N=10,000), monitors demographic parity with real-time alerts (threshold: subgroup intraclass correlation coefficient [ICC] <0.75), and maintains SHA-256 audit trails.

#### Student interface and scaffolding

The EJT presents research outputs to students through a plain-language dashboard designed for grades 9–12 CTE learners. CVET outputs are displayed as a ranked list of community health themes in everyday language (e.g., “Heat illness risk”, “Asthma and air quality”) with color-coded confidence indicators rather than probability scores. ESHI outputs are expressed as “lives protected per year,” “school days recovered,” and “heating, ventilation, and air conditioning (HVAC) cost savings per building,” rather than disability-adjusted life years (DALYs), net present value (NPV), or dose-response coefficients. EJIP outputs are presented as a visual neighborhood investment map with color-coded priority tiers and a plain-language rationale for each ranked investment. All scenario prompts are scaffolded with vocabulary support, worked examples, and sentence starters appropriate for the CTE construction context. Instructors access a parallel facilitator view displaying the same student-facing outputs alongside teaching notes, discussion prompts, and a real-time class progress dashboard. The technical computation layer (dose-response functions, Monte Carlo simulation, equity weighting) operates in the background and is not presented to students during normal use; it is accessible to instructors and researchers through a separate administrative interface.

#### Curriculum structure

Weeks 1–4 cover VDC technical foundations (blueprint reading, building codes, Occupational Safety and Health Administration (OSHA) 10, materials science, carpentry) and are delivered identically in both arms. Weeks 5–10 introduce EJT module training through guided exercises analyzing community interview transcripts. Weeks 11–18 focus on community engagement, including community-based stakeholder interviews and neighborhood health audits. Weeks 19–24 comprise the capstone design phase, in which students integrate VDC drawings, InVEST valuations, and community voice into equity-weighted retrofit proposals.

#### Instructor preparation

CTE instructors assigned to intervention classrooms will complete an 8-hour professional development workshop led by the research team covering: EJT software operation, 51-theme taxonomy application, facilitation of community engagement activities, and recognition of student distress. All intervention-arm instructors additionally hold current Occupational Safety and Health Administration (OSHA) 10 certification, consistent with the safety content taught in the curriculum. Instructors receive detailed lesson plans, slide decks, and a facilitator guide. Ongoing support includes biweekly check-in calls and an instructor support channel. Control arm instructors receive standard curriculum materials without EJT training to maintain separation.

#### Control condition

Traditional VDC curriculum per California CTE Model Curriculum Standards with equal contact hours but without health equity, environmental justice, or planetary health content. Control participants receive the EJT curriculum after study completion (waitlist control).

#### Contamination mitigation

The EJT curriculum requires active software engagement, guided exercises, and a capstone project—content that cannot be passively absorbed through peer interaction; outcome assessors are blinded to group allocation throughout; and where the same instructor teaches students in both arms, temporal separation across class periods is employed with weekly adherence checklists tracking cross-arm exposure. Target fidelity: ≥80% adherence.

### Outcomes

**Table 1** summarizes each outcome measure, its construct, scoring range, assessment timing, and interpretation.

**Table 1.** Summary of Study Outcomes.

| Outcome | Acronym | Construct | Range | Timing | Interpretation |
| --- | --- | --- | --- | --- | --- |
| Health-Integrated Equity Consciousness Index | HI-ECI | Primary: Composite infrastructure literacy | 0–100 | T0,T1,T2*,T3 | Higher = greater equity consciousness |
| Community Health Impact Recognition Score | CHIRS | Secondary: Scenario-based health pathway recognition | 0–100 | T0,T1,T2,T3 | Higher = better recognition |
| Project Health Impact Assessment Score | P-HIAS | Secondary: Capstone equity integration | 0–100 | T2,T3 | Higher = better integration |
| Health Justice Integration Coefficient | HJIC | Exploratory: natural language processing (NLP)-derived equity reasoning depth | 0–1 | T2,T3 | Higher = deeper integration |
| Reflective Health Complexity Score | RHCS | Exploratory: Linguistic complexity of reasoning | 0–100 | T0,T1,T2,T3 | Higher = more complex reasoning |
| Longitudinal Health Engagement Trajectory | LHET | Exploratory: Attitude change slope | Slope | T0 → T3 | Positive = increasing engagement |

Note: \*T2 (6 months) = primary endpoint. T0=baseline, T1=3 months, T2=6 months, T3=12 months.

#### Primary outcome

Health-Integrated Equity Consciousness Index (HI-ECI). Composite measure of students’ capacity to (1) recognize health inequities in infrastructure conditions (Problem Recognition, 40%), (2) integrate equity into technical decisions (Design Integration, 35%), and (3) advocate for community-informed investment (Advocacy Activation, 25%). Scores range 0–100, computed as: HI-ECI = 0.40 × PR_score + 0.35 × DI_score + 0.25 × AA_score, where PR_score (Problem Recognition), DI_score (Design Integration), and AA_score (Advocacy Activation) are each derived from CVET confidence-weighted theme scores for the corresponding sub-domain, normalized to [0, 100] (see **Technical Supplement Section 2.1.1**). Assessed at baseline (T0), 3 months (T1), 6 months (T2, primary endpoint), and 12 months (T3) (see **Table 1**). Scenario prompts present students with a description of a hypothetical infrastructure project in a Disadvantaged Community and ask them to: (1) identify the health equity implications of the proposed design; (2) propose an equity-weighted design modification drawing on community health priorities; and (3) articulate an advocacy strategy for communicating those priorities to relevant stakeholders. Prompts are scaffolded with vocabulary support and worked examples appropriate for grades 9–12 CTE learners; full prompt text and scoring rubrics are pre-registered on ClinicalTrials.gov and available in the study operations manual.

#### Psychometric validation

A two-phase validation protocol applies. Pilot phase (n=52): exploratory factor analysis with oblique rotation evaluates three-factor structure; item-total correlations ≥0.30 and factor loadings ≥0.40 are retention thresholds. Go/No-Go criteria: Cronbach’s α ≥0.70, test-retest r ≥0.75 (2-week interval), inter-rater ICC ≥0.80. Main trial (N=200): confirmatory factor analysis tests the three-factor structure with measurement invariance across school sites (configural, metric, scalar). Convergent validity: correlation with Environmental Justice Literacy Assessment (target r ≥0.50). Discriminant validity: correlation with academic self-efficacy (target r <0.30).

#### Secondary outcomes (see **Table 1**)

(1) Community Health Impact Recognition Score (CHIRS)— scenario-based assessment across 8 health domains (air quality, heat, flooding, noise, mental health, physical activity, food security, housing); each domain scored 0–12.5 points by trained assessors using a domain-specific rubric, summed to a 0–100 composite. (2) Project Health Impact Assessment Score (P-HIAS)—rubric-based capstone evaluation scored across five dimensions: equity integration, technical accuracy, community voice incorporation, feasibility analysis, and advocacy framing (0–20 points each; 0–100 total). Secondary outcomes use Benjamini-Hochberg false discovery rate (FDR) correction at 5%.

#### Exploratory outcomes (see **Table 1**)

(1) Health Justice Integration Coefficient (HJIC)— computed by CVET as the proportion of health equity themes identified in a student response relative to the maximum possible theme set, confidence-weighted (range 0–1; higher = greater integration depth). (2) Reflective Health Complexity Score (RHCS)—Coh-Metrix indices of syntactic complexity and semantic cohesion applied to written responses, scaled to 0–100 (higher = more sophisticated health equity reasoning). (3) Longitudinal Health Engagement Trajectory (LHET)—ordinary least squares slope of HI-ECI scores across T0–T3 (positive slope = increasing engagement over time; a slope of ≥0.5 HI-ECI units per month is pre-specified as the minimum educationally important trajectory, equivalent to a 3-point gain over the 6-month intervention period, consistent with the Go/No-Go progression criterion applied in the pilot phase).

#### Process outcomes

Process measures include implementation fidelity (adherence checklist, completion rate), participant engagement (EJT usage logs), participant satisfaction (validated survey), and adverse event documentation.

### Participant timeline

**Table 2** presents the schedule of enrollment, interventions, and assessments following SPIRIT guidelines. **Figure 1** presents the CONSORT flow diagram for participant enrollment, randomization, follow-up, and analysis. The 12-month total commitment (6-month intervention plus 6-month follow-up) is standard for educational randomized controlled trials (RCTs) assessing sustained knowledge transfer and behavior change, and is consistent with protocols published in BMJ Open and comparable journals. The follow-up period was selected to assess sustainability of educational gains beyond immediate post-intervention and to capture whether health equity consciousness translates into observable behavioral changes during the academic year. The intervention is delivered during regular CTE class periods, requiring no time commitment beyond normal school attendance. Follow-up assessments at T1, T2, and T3 each require approximately 45 minutes and are scheduled during class periods in coordination with participating instructors.

**Figure 1.**
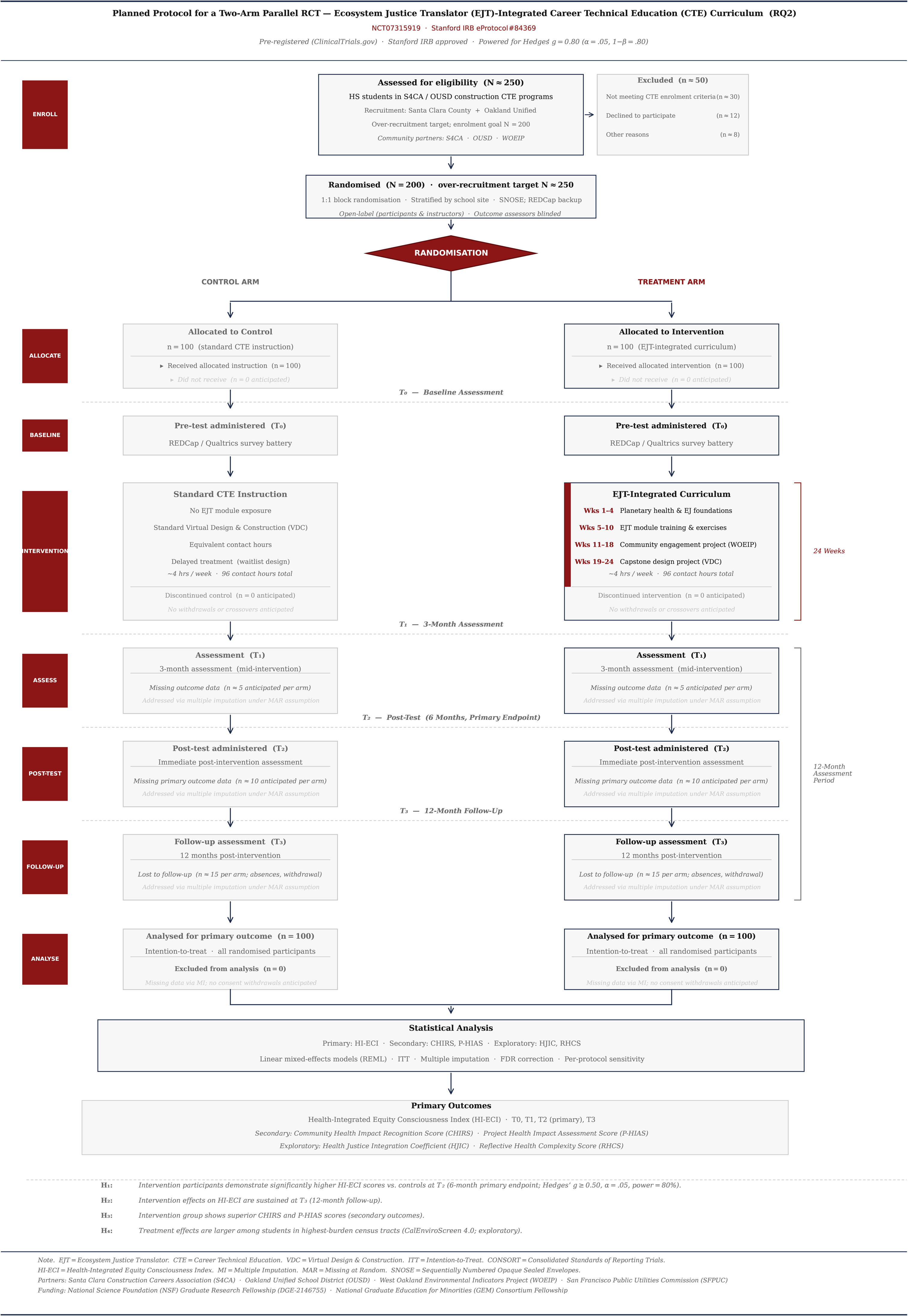
CONSORT flow diagram. Flow diagram illustrating participant enrollment, individual randomization (1:1 allocation to Ecosystem Justice Translator intervention or waitlist control), follow-up assessments at T0 (baseline), T1 (3 months), T2 (6 months, primary endpoint), and T3 (12 months), and inclusion in the intention-to-treat analysis. Enrolled N=200 (n=100 per arm); over-recruitment target N=250.

**Table 2.** SPIRIT Figure: Schedule of enrollment, interventions, and assessments. [CONSORT flow diagram — to be uploaded as a separate high-resolution figure file (minimum 300 dpi, TIFF or PDF format) per BMJ Open figure preparation guidelines]

| TIMEPOINT | Enroll | T0 (0) | T1 (3m) | T2 (6m) | T3 (12m) | Ongoing |
| --- | --- | --- | --- | --- | --- | --- |
| **ENROLLMENT** |  |  |  |  |  |  |
| Eligibility screen | X |  |  |  |  |  |
| Informed consent/assent | X |  |  |  |  |  |
| Allocation | X |  |  |  |  |  |
| **INTERVENTIONS** |  |  |  |  |  |  |
| EJT-integrated curriculum |  | ← — | — | → — |  |  |
| Control (VDC) curriculum |  | ← — | — | → — |  |  |
| **ASSESSMENTS** |  |  |  |  |  |  |
| Demographics |  | X |  |  |  |  |
| HI-ECI (primary) |  | X | X | X* | X |  |
| CHIRS, P-HIAS (secondary) |  | X | X | X | X |  |
| HJIC, RHCS, LHET (exploratory) |  | X | X | X | X |  |
| Process measures |  |  | X | X |  |  |
| Adverse events |  |  |  |  |  | X |

Note: \*T2 (6 months) is the primary endpoint.

### Sample size

Sample size was calculated using G*Power 3.1.9.7 (Faul et al., 2007)^26^: independent samples t-test, two-tailed α=0.05, power (1−β)=0.80, anticipated Hedges’ g=0.80, calibrated to meta-analyses of active learning in science, technology, engineering, and mathematics (STEM) (g=0.80+; Freeman et al., 2014)^27^ and health education among vulnerable youth (g=0.70–0.90; Gefter et al., 2021).^28^

Sensitivity analyses confirm adequate power (>70%) to detect Hedges’ g=0.70 with n=85 per group retained after anticipated attrition. Effect sizes are reported as Hedges’ g with small-sample J bias-correction (J=1−3/(4(N−2)−1); for N=200, J≈0.996; Hedges, 1981).

### Recruitment

Participants will be recruited through: (1) presentations during CTE classes; (2) information sheets via school email and backpack mail; (3) CTE coordinator referrals; (4) follow-up calls to families. Recruitment continues until target achieved or 8 weeks maximum. Participant incentives are for student participants only: $25 at baseline (T0), $25 at T2 (6 months), and $50 at T3 (12 months), for a maximum of $100 for full protocol completion. Students completing only baseline receive $25; students completing baseline and T2 receive $50 total; students completing all three timepoints receive $100 total. Post-randomization withdrawals retain incentives earned to date and are not replaced. No financial incentives are provided to CTE instructors; instructor participation is framed as a professional development activity within their normal teaching role, and participating instructors will receive a letter of recognition suitable for professional development portfolios upon study completion. This non-financial instructor acknowledgment was determined to be appropriate and non-coercive by the Stanford IRB.

### Randomization and allocation concealment

#### Sequence generation

The allocation sequence will be computer-generated using R statistical software (blockrand package) with block randomization using randomly varying block sizes (4, 6, 8) stratified by school site. The sequence will be generated by an independent biostatistician at Stanford who is not involved in participant enrollment or outcome assessment.

#### Allocation concealment

Sequentially numbered, opaque, sealed envelopes (SNOSE) prepared by the independent biostatistician. Envelopes are stored securely and opened only after participant enrollment has been confirmed and baseline assessment completed. REDCap serves as a backup randomization system.

#### Blinding

The trial is open-label for participants and instructors, as blinding is not feasible in educational intervention research. Outcome assessors are blinded via: (1) de-identified materials submitted under study IDs only; (2) a separate assessment team with no knowledge of group assignment. The study statistician is blinded using coded group identifiers (A/B).

### Data collection and management

Data are collected via REDCap (Health Insurance Portability and Accountability Act [HIPAA]-compliant, Stanford-hosted), with Qualtrics (Qualtrics XM Platform, Provo, UT) used alongside REDCap for data logging. Demographics, outcomes, and process measures are collected at each scheduled timepoint. EJT usage is logged automatically. Qualtrics log entries are linked to REDCap records via randomly generated study ID, with no participant identifiers retained within Qualtrics. All data are de-identified; linking files are stored separately with restricted access. The system employs Advanced Encryption Standard 256-bit (AES-256) encryption, two-factor authentication, and monthly quality audits. EJT software is deployed as a Progressive Web Application (PWA) compatible with all major browsers (Chrome, Firefox, Safari, Edge) and functional on Chromebook devices (the predominant school-issued device in OUSD schools), iOS, and Android, without software installation. Compatibility with OUSD content filtering and firewall settings will be confirmed through a pre-study technical assessment conducted with each school’s technology coordinator prior to the start of data collection. IP addresses captured in REDCap server logs are automatically excluded from research data exports; access logs are retained separately under restricted access per Stanford University IT Security Policy and are not linked to participant records. All data transmissions use Transport Layer Security (TLS) 1.3 encryption. No student identification numbers, names, or school identifiers are transmitted to external servers; participants are identified only by randomly generated study IDs throughout all data systems.

### Statistical methods

#### Primary analysis

Intention-to-treat. Linear mixed-effects models using restricted maximum likelihood (REML) with random intercepts for participants (within-person correlation across repeated assessments); fixed effects for treatment group (intervention vs. control), time (categorical: T0, T1, T2, T3), treatment×time interaction (primary test), school site, and baseline HI-ECI score. Primary test: treatment effect at T2 (6 months), two-sided p<0.05, 95% CI. Effect sizes: Hedges’ g with small-sample J bias-correction (J≈0.996 for N=200; Hedges, 1981).

#### School site as fixed effect

Fixed (not random) because the anticipated small number of sites provides insufficient degrees of freedom for reliable random effects variance estimation. Controls for systematic between-school differences in demographic composition, baseline academic performance, instructor experience, and facilities.

#### Secondary analyses

Heterogeneous treatment effects are examined via interaction terms (CalEnviroScreen quartile, sex, race/ethnicity, baseline HI-ECI). Benjamini-Hochberg FDR correction at 5% is applied for all secondary outcomes. A per-protocol sensitivity analysis is conducted for participants with ≥80% intervention attendance.

#### Missing data

Missing data are assessed using Little’s MCAR test. The primary analysis assumes data are missing at random (MAR); linear mixed-effects models use all available data under this assumption. Sensitivity analyses include: (1) multiple imputation by chained equations (MICE) with 50 imputations; (2) pattern-mixture models under missing not at random (MNAR); and (3) tipping-point analysis. Differential dropout by arm is assessed via Fisher’s exact test if attrition exceeds 10% per arm. Analyses use R 4.3+ (lme4, mice, emmeans packages).

### Data monitoring and safety

#### Data monitoring

No formal Data Safety Monitoring Board (DSMB) is required given the minimal-risk educational intervention design (per Stanford IRB guidance for minimal-risk educational research). The principal investigator (PI) serves as safety monitor reviewing enrollment, retention, adverse events, and deviations monthly. An interim safety report will be submitted to the IRB at 3 months. No interim efficacy analyses are planned.

#### Adverse events

Anticipated risks are minimal and limited primarily to potential distress arising from discussions of environmental injustice in students’ own communities. Instructors are trained to recognize signs of distress and to provide referrals to school counseling services. All adverse events are documented with description, severity, relationship to the study, and resolution. Serious adverse events are reported to the Stanford IRB within 24 hours of identification. Pre-specified stopping rules are: (1) two or more serious adverse events judged related to study participation; (2) dropout exceeding 50% per arm; or (3) IRB request to halt the trial.

## Supporting information

Technical Supplementary File

SPIRIT Checklist File

## ETHICS AND DISSEMINATION

### Ethics approval

This protocol has been approved by the Stanford University Institutional Review Board (IRB eProtocol #84369, approved February 13, 2026). OUSD research committee approval will be obtained prior to recruitment. The trial is registered on ClinicalTrials.gov (NCT07315919). Protocol amendments will be submitted to the IRB, updated on the trial registry, and communicated to all participants.

### Consent

Written parental consent and student assent obtained by research coordinators (not instructors) in private settings. Consent forms are available in English and Spanish. Participants may withdraw from the study at any time without penalty or loss of any incentives earned to date.

### Confidentiality

All participant data are de-identified using randomly generated study IDs. Data are stored on encrypted Stanford servers with restricted access controls. Paper documents are kept in locked storage. Data retention and destruction follow Stanford University Research Policy 4.1: all study records retained for a minimum of seven years following study completion or last participant contact, whichever is later, per Stanford IRB requirements for studies involving minors. Consent forms and identifying linkage files are stored separately from de-identified research data in locked, secure locations accessible only to authorized study personnel. Adverse event records are retained for the duration required by applicable regulations. These retention requirements are documented in the IRB-approved protocol (eProtocol #84369) and will not be modified without IRB approval. All data collection and storage procedures are compliant with the Family Educational Rights and Privacy Act (FERPA).

### Post-trial care

Control arm participants receive the full EJT-integrated curriculum following study completion (waitlist design). All participants receive a certificate of completion acknowledging their contribution to the research.

### Replicability

All instruments, rubrics, curriculum, and EJT source code will be released under CC BY-NC-SA 4.0 concurrent with primary publication, permitting free use for educational, research, and non-profit purposes. Commercial licensing may be pursued separately through Stanford University Office of Technology Licensing (OTL docket S25-565). The version-locked large language model specification is documented on ClinicalTrials.gov (NCT07315919). De-identified data and code available on OSF upon request. A detailed implementation toolkit will be made freely available for educators and researchers concurrent with primary publication.

### Dissemination

Results will be disseminated in peer-reviewed open-access journals regardless of outcome, targeting BMJ Open (protocol), Lancet Planetary Health (results), and the Journal of Vocational Education and Training. This protocol will also be posted as a preprint on medRxiv concurrent with journal submission to enable early, open access ahead of peer review. Findings will be presented at the American Educational Research Association (AERA) and American Public Health Association (APHA) conferences. Community-first dissemination will ensure findings reach community partners and Oakland families before journal submission, pending formal agreements. Authorship follows International Committee of Medical Journal Editors (ICMJE) criteria. Publication of this protocol prior to data collection follows established best practices for randomized trial transparency endorsed by SPIRIT 2013 guidelines, ICMJE requirements, and BMJ Open editorial policy. This approach enables independent verification that reported analyses match pre-specified plans, reducing risk of selective outcome reporting bias.

### Patient and public involvement

The EJT 51-theme taxonomy was derived from validated public health frameworks (CDC SVI, EPA EJScreen, Healthy People 2030). Anticipated collaboration with WOEIP and the Santa Clara County Construction Careers Association (S4CA), pending formal agreements, would support: (1) classroom access and participant recruitment; (2) curriculum review with S4CA and OUSD instructors; (3) community liaison and stakeholder engagement. Upon agreement, WOEIP would hold veto rights over misrepresentative interpretations. Community-first dissemination would ensure findings reach Oakland families before publication. Intervention burden was assessed through feedback from student advisors during the curriculum development phase.

## AUTHOR CONTRIBUTIONS

Devan Cantrell Addison-Turner: Conceptualization, Methodology, Software, Formal analysis, Investigation, Writing—original draft, Project administration, Funding acquisition. Gretchen Cara Daily: Conceptualization, Methodology, Supervision, Resources, Writing—review/editing, Funding acquisition. Both approved final manuscript.

## DATA AVAILABILITY STATEMENT

Protocol paper; no data yet available. Following completion, de-identified data and code available on OSF upon request with data use agreement. EJT software and curriculum on GitHub concurrent with publication.

## ACKNOWLEDGEMENTS

We thank WOEIP (Ms. Margaret Gordon, Board Chair; Ms. Veronica Eady, Interim Executive Director) for expressing interest in community collaboration. We acknowledge S4CA (Dr. Brenda Childress, Executive Director) and OUSD CTE coordinators for their expressed interest in supporting this research. Protocol developed as part of Devan Cantrell Addison-Turner’s doctoral research, Department of Civil and Environmental Engineering, Stanford Doerr School of Sustainability, conducted within the Natural Capital Alliance, Lepech Research Group, and Center for Integrated Facility Engineering. The authors thank Kristy Hsiao for generous individual support of this research.

