## Supplementary material for "Integrating planetary health and environmental justice into high school construction career education: protocol for a randomized controlled trial of the Ecosystem Justice Translator": Technical Supplementary File

### Technical Supplement

#### Ecosystem Justice Translator (EJT) v4.1

*Cyber-Physical Control System for Health Equity Investment Optimization*

Supplementary File 2 for BMJ Open Protocol

Devan Cantrell Addison-Turner[1], Gretchen C. Daily[2]

*1. Stanford University, Civil and Environmental Engineering, Stanford, CA 94305, USA | 2. Stanford University, Stanford, CA 94305, USA | ORCID: Devan Cantrell Addison-Turner 0000-0002-2511-3680; Gretchen C. Daily 0000-0003-1443-1111*

Protocol ID: Institutional Review Board (IRB)-84369 | ClinicalTrials.gov NCT07315919 | June 2026

**ABBREVIATIONS:** AHP, Analytic Hierarchy Process; ATSDR, Agency for Toxic Substances and Disease Registry; CDC, Centers for Disease Control and Prevention; CVET, Community Voice Equity Translation; DALY, Disability-Adjusted Life Year; DP, Demographic Parity; EJT, Ecosystem Justice Translator; EJIP, Environmental Justice Investment Prioritization; EPA, Environmental Protection Agency; ESHI, Ecosystem Service Health Integration; ICC, Intraclass Correlation Coefficient; InVEST, Integrated Valuation of Ecosystem Services and Tradeoffs; IRB, Institutional Review Board; LLM, Large Language Model; NER, Named Entity Recognition; PWA, Progressive Web App; ROI, Return on Investment; SUS, System Usability Scale; SVI, Social Vulnerability Index; UBR, Uncertainty, Bias, and Risk; WPM, Weighted Product Model; ECE, Expected Calibration Error

### 1. SYSTEM OVERVIEW

The Ecosystem Justice Translator (EJT) v4.1 is a **cyber-physical control system** implementing multi-dimensional optimization across health equity outcomes (Disability-Adjusted Life Years [DALYs]), financial returns (Return on Investment [ROI] and payback period), and time savings (recovered instructional days and workforce hours), as defined in Sections 4.1.1–4.1.2. The system uses equity weights derived from vulnerability indices to ensure Pareto-optimal investment decisions that prioritize Disadvantaged Communities.

**Core Innovation:** EJT bridges the gap between community voice (qualitative) and investment optimization (quantitative) by using Large Language Models (LLMs) to extract structured health equity themes from unstructured community input, then linking these themes to validated epidemiological dose-response functions for quantified health benefit estimation.

#### 1.1 Key System Features

##### Table 1. EJT Key System Features

| Feature | Description |
|---|---|
| **Multi-Objective Optimization** | Simultaneous optimization across health equity outcomes (DALYs), financial returns (ROI, NPV, payback period), and time savings (recovered instructional days) |
| **Equity-Weighted Prioritization** | Centers for Disease Control and Prevention (CDC) Social Vulnerability Index (SVI) amplifies benefits for disadvantaged communities |
| **In-Browser Machine Learning** | TensorFlow.js enables local processing; no data leaves user device |
| **Progressive Web App (PWA)** | Offline capability, mobile-responsive, installable application |
| **3+3 Validation Framework** | 3 primary validation metrics + 3 supporting system checks |
| **Student-Facing Interface** | Plain-language dashboard for grades 9–12 learners: health themes in everyday language, outputs expressed as lives protected, school days recovered, and cost savings; neighborhood investment map with color-coded priority tiers; scaffolded scenario prompts with vocabulary support. Parallel instructor facilitator view with teaching notes and class progress dashboard. Technical computation layer (dose-response, Monte Carlo, equity weighting) runs in background; not presented to students during normal use. |

#### 1.2 System Architecture — Four Integrated Modules

##### Table 2. EJT System Architecture — Four Integrated Modules

| Module | Function | Key Technologies |
|---|---|---|

| | | |
|---|---|---|
| **1. CVET** | Community Voice Equity Translation | Most recent generally available Claude model (Anthropic; version-locked prior to enrollment) with 51-theme health equity taxonomy |
| **2. ESHI** | Ecosystem Service Health Integration | Integrated Valuation of Ecosystem Services and Tradeoffs (InVEST) + epidemiological dose-response functions |
| **3. EJIP** | Environmental Justice Investment Prioritization | Multi-criteria optimization with Analytic Hierarchy Process (AHP) + Monte Carlo |
| **4. UBR** | Uncertainty, Bias, and Risk Quantification | Monte Carlo (N=10,000), demographic parity monitoring |

### 2. MODULE SPECIFICATIONS

#### 2.1 Community Voice Equity Translation (CVET)

**Purpose:** Extract structured health equity themes from unstructured community input (interview transcripts, survey responses, public comments, social media) using LLMs.

**Theme Extraction Algorithm:**

1. Tokenize document *d* into segments $S = \{s_1, s_2, ..., s_m\}$
2. For each segment $s_j$: Generate embedding $e(s_j)$ using fine-tuned transformer encoder
3. Compute theme logits: $L(s_j) = W \times e(s_j) + b$, where W is theme classification matrix
4. Apply temperature-scaled softmax: $P(t_i|s_j) = \exp(L_i/T) / \Sigma_k \exp(L_k/T)$
5. Extract named entities for geographic attribution using Named Entity Recognition (NER): $G(s_j) = NER(s_j)$
6. Aggregate segment-level themes to document-level with confidence weighting

##### 2.1.1 CVET Fine-Tuning and Prompt Engineering Specification

**Implementation approach:** The CVET module implements the 51-theme health equity taxonomy through structured prompt engineering applied to the most recent generally available Claude model (Anthropic) at the time of trial commencement, rather than weight-level fine-tuning of the base model. This approach was selected for three reasons: (1) prompt engineering preserves the full generative and reasoning capabilities of the base model while enabling domain-specific theme classification; (2) it avoids the need for large labeled training corpora that would require additional data use agreements with community partners; and (3) it enables exact reproducibility through version-locking the model snapshot and archiving the system prompt, without requiring access to proprietary fine-tuning infrastructure.

**System prompt structure:** The CVET system prompt comprises four components: (1) a role specification instructing the model to act as a health equity theme classifier trained on the 51-theme taxonomy; (2) the full 51-theme taxonomy with domain assignments, operational definitions, and example phrases for each theme; (3) output format specification requiring structured JSON with theme identifier, probability score $\in [0,1]$, confidence tier (high/medium/low), and supporting text span; and (4) calibration instructions specifying temperature parameter T=0.2 for reduced output variance and instructions for handling ambiguous or multi-theme segments. The complete system prompt is version-locked, SHA-256 hashed, and archived on the Open Science Framework (OSF) concurrent with trial commencement, enabling exact replication.

**Temperature calibration:** The temperature-scaled softmax in Step 4 of the theme extraction algorithm (Section 2.1) applies a calibration temperature T derived from held-out validation data using Platt scaling. Calibration was performed on a development set of 200 manually annotated community interview segments (not drawn from study participants), with theme labels assigned by two independent coders (Cohen's $\kappa \geq 0.75$ prior to adjudication). The calibrated temperature minimizes Expected Calibration Error (ECE) on the held-out set. The calibration dataset, annotation codebook, and inter-rater reliability statistics are archived on OSF under restricted access (available to researchers on request with data use agreement).

**Version locking and reproducibility:** The CVET module uses the most recent generally available Claude model (Anthropic) at the time of trial commencement; this snapshot is then pinned via the Anthropic API using the 'anthropic-version' header and a fixed model string, ensuring all participant responses are processed through identical model weights throughout the trial period. The selected model string, together with API parameters (temperature, max_tokens, top_p), system prompt SHA-256 hash, and API call logs, are recorded in the SHA-256 audit trail maintained by the UBR module (Section 4.2). In the event that Anthropic deprecates the pinned model version during the trial, the research team will: (1) immediately notify the IRB; (2) process all remaining responses through a documented successor model with a sensitivity analysis comparing outputs across model versions on a bridging sample; and (3) report any model transition transparently in the primary outcomes paper.

**Application to capstone projects:** During the capstone design phase (Weeks 19–24), CVET processes three categories of student-generated text through the same theme extraction pipeline: (1) community interview transcripts collected during Weeks 11–18, which generate the 'community voice' input to the EJIP formula; (2) written advocacy strategy sections of the capstone proposal, which contribute to the Advocacy Activation sub-domain of HI-ECI; and (3) design rationale narratives, which contribute to the Design Integration sub-domain. The ESHI module concurrently processes the green-blue infrastructure investment specified in the student's VDC drawings, translating the proposed intervention (tree canopy area, green roof footprint, bioswale capacity) through the dose-response library into plain-language health benefit estimates (lives protected per year, school days recovered, HVAC cost savings). The EJIP module then combines CVET community voice scores, ESHI health benefit estimates, census-tract SVI equity weighting, and a feasibility score into a ranked investment priority that forms the analytical spine of the capstone proposal. Students interact with these outputs exclusively through the plain-language dashboard; the underlying LLM calls are not visible during normal student use.

**51-Theme Health Equity Taxonomy (8 Domains)**

**Table 3. 51-Theme Health Equity Taxonomy by Domain**

| Domain | Themes | Example Themes |
|---|---|---|
| Environmental Exposures | 8 | Air quality index exceedance; Extreme heat exposure; Flood and stormwater risk; Noise pollution; Proximity to hazardous waste sites; Pesticide exposure; Greenspace access deficit; Indoor air quality |
| Health Outcomes | 7 | Respiratory illness burden; Cardiovascular disease risk; Mental health and depression; Heat illness and mortality; Childhood asthma prevalence; Waterborne disease exposure; Physical activity and obesity |
| Social Determinants | 9 | Housing quality and overcrowding; Employment and economic stability; Transportation access and burden; Food security and desert status; Educational attainment; Language and social isolation; Healthcare access; Poverty and income inequality; Disability status and vulnerability |
| Procedural Justice | 6 | Community participation in planning decisions; Government transparency and information access; Accountability mechanisms for decision-makers; Meaningful access to decision-making processes; Notification and early community engagement; Anti-displacement protections |
| Distributive Justice | 5 | Equitable resource allocation across communities; Fair distribution of environmental benefits; Proportionate sharing of environmental burdens; Investment prioritization for disadvantaged communities; Benefit-cost distribution across income levels |
| Recognition Justice | 6 | Cultural respect and identity affirmation; Historical acknowledgment of environmental harm; Community self-determination rights; Traditional ecological knowledge recognition; Anti-racism in environmental planning; Intersectional vulnerability acknowledgment |
| Restorative Justice | 4 | Environmental remediation of contaminated sites; Community compensation and reparative investment; Healing-centered green infrastructure; Reconciliation processes with affected communities |
| Community Assets | 6 | Accessible green space and parks; Social cohesion and community trust; Local economy and business resilience; Cultural heritage sites and community spaces; Community-led health initiatives; Neighborhood safety and violence prevention |
| **TOTAL** | **51** | *Derived from CDC SVI, Environmental Protection Agency (EPA) EJScreen, Healthy People 2030* |

##### 2.1.2 HI-ECI Scoring Pipeline

**Purpose:** Convert a student's written response to a standardized scenario prompt into a sub-domain score for each of the three HI-ECI dimensions (Problem Recognition, Design Integration, Advocacy Activation), then aggregate into the 0–100 composite HI-ECI score.

**Step 1 — Response Segmentation and Theme Extraction:** Each student response d is tokenized and processed through the CVET theme extraction algorithm (Section 2.1 steps 1–6). This yields a document-level theme probability vector $P(t_i|d) \in [0,1]$ for each of the 51 themes, with confidence score $c_i$ derived from temperature-scaled softmax.

**Step 2 — Sub-Domain Score Computation:** Each of the 51 themes is pre-assigned to one of the three HI-ECI sub-domains by the instrument development team. The raw sub-domain score for sub-domain k is:

$RawScore\_k(d) = [\Sigma_i \in T\_k \ (P(t_i|d) \times c_i)] / |T\_k|$

Where T_k is the set of themes assigned to sub-domain k, $P(t_i|d)$ is the CVET-derived theme probability for theme i in response d, $c_i$ is the confidence score for that theme assignment, and |T_k| is the cardinality of T_k (number of themes in sub-domain k). RawScore_k(d) ∈ [0,1]. Each sub-domain score is then normalized to [0, 100]: Score_k(d) = RawScore_k(d) × 100.

**Step 3 — HI-ECI Composite Score:** Sub-domain scores are aggregated using pre-specified weights:

HI-ECI(d) = 0.40 × Score_PR(d) + 0.35 × Score_DI(d) + 0.25 × Score_AA(d)

Where Score_PR = Problem Recognition sub-domain score (themes addressing recognition of infrastructure health inequities), Score_DI = Design Integration sub-domain score (themes addressing equity-weighted technical design decisions), and Score_AA = Advocacy Activation sub-domain score (themes addressing community-centred investment advocacy). Sub-domain theme assignments and weights are fixed prior to data collection and documented on ClinicalTrials.gov (NCT07315919).

**Step 4 — Inter-Rater Reliability Verification:** A random 20% sample of responses is independently scored by a second trained assessor blind to the CVET output. Agreement is assessed using intraclass correlation coefficient (ICC; two-way mixed, absolute agreement). Target ICC ≥ 0.80. Where ICC falls below threshold, responses are adjudicated by a third assessor and the CVET confidence threshold is recalibrated before proceeding to primary analysis.

#### 2.2 Ecosystem Service Health Integration (ESHI)

Purpose: Quantify health benefits of green-blue infrastructure investments using validated ecosystem service models (InVEST)[1] linked to epidemiological dose-response functions from peer-reviewed meta-analyses.

**Technical Implementation:**

- Ecosystem Service Modeling: InVEST v3.12
- Spatial Resolution: 30m grid cells (aligned with Landsat imagery)
- InVEST Models: Urban Cooling, Urban Stormwater Retention, Recreation Opportunity
- Vulnerability Weighting: CDC/Agency for Toxic Substances and Disease Registry (ATSDR) SVI at census tract level

**InVEST-to-dose-response pipeline:** InVEST model outputs (biophysical quantities: cooling energy in MWh/year, runoff retention in $m^3$/year, recreation access in person-visits/year) are translated into epidemiological exposure variables at the census-tract level through a three-step pipeline. First, biophysical outputs are spatially aggregated from 30m grid cells to census-tract polygons using area-weighted averaging, preserving proportional exposure at the population level. Second, exposure variables are converted to the units required by each dose-response function: urban cooling output (ΔT in °C) feeds the Gasparrini heat mortality function; $PM_{2.5}$ mass removal (Δμg/$m^3$) feeds the Hoek cardiorespiratory mortality function; greenspace access area (Δha within 300m) feeds the Gascon mental health and depression functions. Third, population-level health outcomes are computed by multiplying dose-response effect estimates by census-tract population counts, stratified by age and vulnerability group using CDC/ATSDR SVI weights. Uncertainty in InVEST biophysical outputs (arising from input data precision and model parameter uncertainty) is treated as a fixed scaling uncertainty of ±15% on all biophysical quantities, propagated through the Monte Carlo simulation in UBR (Section 4.2) by sampling uniformly over this range.

### 3. DOSE-RESPONSE FUNCTION LIBRARY

The EJT dose-response function library contains ten peer-reviewed exposure-response relationships linking green-blue infrastructure outcomes to quantified health endpoints. Each function was selected on three criteria: (1) derivation from a systematic review or meta-analysis rather than a single primary study; (2) reported effect estimates compatible with the spatial resolution of InVEST model outputs (census-tract or neighborhood level); and (3) applicability to the urban California context in which the trial is conducted. Where a function was derived from non-U.S. populations[2,3], sensitivity analyses using California-specific effect estimates are pre-specified in the analysis plan. The library is extensible: additional functions may be added through the EJT administrative interface by researchers with appropriate credentials, with each addition logged in the UBR audit trail (Section 4.2) and requiring re-validation against the 3+3 framework (Section 5) before use in primary analyses. **Table 4** presents the complete dose-response function library with effect sizes, sources, and applicability conditions.

**Table 4. Dose-Response Function Library**

| Health Outcome | Function | Effect Size | Source |
|---|---|---|---|
| Heat mortality | ΔM = 0.0387 × ΔT × Pop × BR | 3.9% per 1°C (see note a) | [2] |
| PM2.5 mortality | HR = 1.06 per 10 μg/m³ | 1.06% per 10 μg/m³ | [4] |
| Mental health (DALY) | ΔDALY = 0.12 × ΔAccess × Pop | 0.12 DALYs/ha | [3] |
| Depression odds | OR = 0.72 per ha within 300m | 28% lower odds | [3] |
| Nature access (wellbeing threshold) | ≥120 min/week outdoors | Good health and wellbeing | [5] |
| Childhood asthma | OR per 10% canopy increase | 24% lower prevalence | [6] |
| Stress (cortisol) | Δσ = -0.21 × NatureMin | 21% reduction | [7] |
| Energy savings | $4,800/yr per 1,000 trees | 15-30% HVAC reduction | [8] |
| Flood injury (stormwater retention) | Risk reduction per 100m³ retention | Injury/mortality reduction | [9] |
| Noise exposure—reading comprehension (children, chronic traffic/aircraft noise) | OR = 1.31 per 5 dB(A) increase in chronic noise exposure | 31% higher odds of reading impairment per 5 dB(A) | [10] |

Note a: Effect size of 3.9% per 1°C is rounded[2] for presentation consistency; full precision value (3.87%) available in the primary source. Sensitivity analyses using California-specific mortality displacement estimates are pre-specified in the study analysis plan.

### 4. EQUITY WEIGHTING METHODOLOGY

The EJT applies equity weighting to amplify benefits for Disadvantaged Communities. This approach is grounded in published environmental justice scholarship demonstrating that historical patterns of disproportionate environmental burden in low-income and minority communities require affirmative investment prioritization to achieve equitable outcomes.[11,12,13] Equity weighting is further supported by federal legislation including the Inflation Reduction Act (Pub. L. 117-169) and the Biden Administration's Justice40 Initiative (Executive Order 14008, 2021), which directed that 40% of federal climate and clean energy investment benefits flow to Disadvantaged Communities. The trial's equity weighting methodology is independent of any specific regulatory program: the CDC Social Vulnerability Index (SVI 2022) data underlying the weighting algorithm were downloaded and archived locally prior to December 2025. The α parameter is user-adjustable (range [0, 2]; default 0.5) to allow sensitivity analyses with no equity amplification (α = 0).

**SVI-Weighted Benefit Calculation:**

$$\mathbf{HB\_weighted(u) = HB\_raw(u) \times (1 + \alpha \times SVI(u))}$$

**Where:**

- $HB\_raw(u) = \Sigma_j\ DRF_j(ES\_output_j(u))$ summed over all ecosystem services j
- α = Equity amplification parameter (default = 0.5; range [0, 2])
- SVI(u) = CDC/ATSDR Social Vulnerability Index ∈ [0, 1]

**Example:** A census tract with SVI = 90th percentile (0.90) receives weight multiplier of 1 + (0.5 × 0.90) = **1.45×** benefit weight in the prioritization algorithm.

#### 4.1 Environmental Justice Investment Prioritization (EJIP)

**Purpose:** Generate ranked investment priorities using multi-criteria optimization (Weighted Product Model with AHP) that integrates community voice, health benefits, equity weighting, and implementation feasibility.

**Priority Score Formula:**

$$\mathbf{Priority(u) = w_1{\times}Voice(u) + w_2{\times}Health(u) + w_3{\times}Equity(u) + w_4{\times}Feasibility(u)}$$

**Decision-theoretic basis and Pareto-optimality:** The weighted product model (WPM) used in EJIP approximates Pareto-optimality under the condition that all sub-component scores are normalized to [0, 1] and weights sum to 1.0. Under these conditions, any solution that maximizes Priority(u) is Pareto-optimal with respect to the four sub-components: no alternative census tract can improve on any one dimension without reducing at least one other. The WPM was selected over the Analytic Hierarchy Process (AHP) pairwise comparison method because it avoids rank reversal under normalization — a known instability when the number of alternatives (census tracts) is large relative to the number of criteria. The Analytic Hierarchy Process is retained for weight elicitation from community stakeholders; the WPM applies those elicited weights in the aggregation step. This separation of weight elicitation (AHP) from weight application (WPM) is consistent with the multi-criteria decision analysis literature [14] and is documented in the analysis plan pre-registered on ClinicalTrials.gov (NCT07315919). Monte Carlo sensitivity analyses (Section 4.2) test the robustness of the top-five ranked census tracts to ±20% perturbation of all four weights simultaneously, providing a direct empirical test of rank stability under weight uncertainty.

**Table 5. EJIP Priority Score Components and Default Weights**

| Component | Default Weight | Definition |
|---|---|---|
| Voice(u) — Community alignment | $w_1 = 0.40$ | Average confidence-weighted theme score |
| Health(u) — Health benefit | $w_2 = 0.30$ | Composite of DALY reduction, ROI, and time savings (see Sections 4.1.1–4.1.2); sub-component weights pre-specified in study analysis plan and user-adjustable; each sub-component normalized to [0, 1] before aggregation |
| Equity(u) — Vulnerability | $w_3 = 0.20$ | Direct SVI score (0-1) |
| Feasibility(u) — Implementation | $w_4 = 0.10$ | Land availability, cost, infrastructure |

##### 4.1.1 Financial Benefit Calculation

**Purpose:** Translate epidemiological dose-response outputs into monetized financial returns, enabling cost-effectiveness comparison across green-blue infrastructure investment scenarios.

**Annualized Health Savings:** Health-related financial savings are calculated by monetizing dose-response outputs using validated unit cost parameters:

$S\_health(u) = \Sigma j\ [DRFj(ES\_outputj(u)) \times UnitCostj]$

Where UnitCostj is the monetized unit value per health outcome unit for ecosystem service j. Reference unit costs: heat mortality avoided = $12.4 million per statistical life (U.S. EPA central VSL estimate, 2023 dollars)[15]; $PM_{2.5}$ mortality avoided = $12.4 million per statistical life; flood-related mortality avoided = $12.4 million per statistical life (applied with EPA VSL)[9]; chronic absenteeism day recovered = $65.34 per student per day (California Local Control Funding Formula [LCFF] average daily attendance rate; California Department of Education, 2024); HVAC energy savings = $4,800 per year per 1,000 trees[8]. The VSL carries substantial uncertainty; the EPA reports a 5th–95th percentile range of $1.8–$22.4 million (2023 dollars)[15]. Because this empirical range is asymmetric in log-space, it cannot be exactly reproduced by a single symmetric distribution; for tractability, the Monte Carlo simulation (Section 4.2) models VSL as a stochastic parameter, $VSL \sim LogNormal(\mu = \ln(12.4), \sigma = 0.4)$, which yields a comparable 5th–95th percentile range of $6.4–$23.9 million and preserves the EPA's central estimate and approximate uncertainty magnitude[15].

**Return on Investment (ROI):** $ROI(u) = (S\_health(u) - C\_annualized(u)) / C\_annualized(u) \times 100\%$

**Net Present Value (NPV):** $NPV(u) = \Sigma_t\ [S\_health(u) / (1 + r)^t] - C\_total(u)$

Where t indexes years 1 through T (equipment lifespan; default T = 20 years), r is the social discount rate (default r = 3%, per U.S. Office of Management and Budget Circular A-4, 2023; sensitivity range r = 2%–7% to reflect ongoing methodological debate on long-horizon environmental discounting), and C_total(u) is the total upfront intervention cost for census tract u. The discount rate r is user-adjustable within the EJT interface, consistent with the treatment of other analytical parameters. The payback period (total cost / S_health(u)) represents the year at which NPV = 0 under r = 0 and serves as a discount-rate-free complement to NPV for audiences less familiar with discounted cash flow methods.

Where C_annualized(u) = total intervention cost / T for use in the single-period ROI formula. All financial metrics (ROI, NPV, payback period) are computed per census tract and normalized to [0, 1] for integration into the EJIP Health(u) sub-component.

##### 4.1.2 Time Savings Calculation

**Purpose:** Translate health outcome improvements into recovered instructional days and productive workforce hours, providing a time-domain metric alongside financial and health dimensions in the EJIP prioritization score.

Recovered Instructional Days (school populations): For census tracts containing K–12 schools, chronic absenteeism reduction derived from the greenspace–attendance dose-response relationship (a one-unit increase in NDVI within 250 m of a school is associated with a 2.1-day reduction in annual illness-related absences)[16] is converted to recovered student-days:

$T\_school(u) = N\_students(u) \times \Delta Absenteeism(u) \times 25$

Where N_students(u) is the enrolled student population in census tract u, ΔAbsenteeism(u) is the predicted proportional reduction in chronic absenteeism from green-blue infrastructure investment (derived from ESHI), and 25 is the mean annual absence duration for chronically absent students.[17,18]

Recovered Workforce Hours (industry-adjusted): Extending the time-savings logic above to working-age populations, health-related productivity gains are converted to recovered full-time-equivalent work hours, weighted by each census tract’s industry composition to reflect the differential sensitivity of missed work time to health burden across sectors:

$W(u) = \Delta DALY\_wa(u) \times HoursPerDALY \times \Sigma_i\ [ShareInd_i(u) \times IndFactor_i]$

Where ΔDALY_wa(u) is the working-age-attributable DALY reduction in census tract u — the portion of the tract-level dose-response outputs in Section 3 (which are already population-scaled, e.g. the DALY function in Table 4) attributable to working-age (18–64) residents; HoursPerDALY = 2,080 is a fixed conversion constant translating one averted DALY into recovered full-time-equivalent work hours, consistent with an energy-burden-to-productive-hours framework[19]; $ShareInd_i(u)$ is the share of tract u's working-age labor force (U.S. Census Bureau American Community Survey 5-year estimates) employed in industry sector i (U.S. Bureau of Labor Statistics Quarterly Census of Employment and Wages, allocated to census tracts via ZIP-code crosswalk); and $IndFactor_i$ is a sector-specific elasticity coefficient scaling the DALY-to-hours conversion by each industry's differential sensitivity to health-related productivity loss, calibrated as each sector's 2024 national nonfatal occupational injury and illness incidence rate normalized to the highest-incidence sector (transportation and warehousing = 1.00 [reference category, 4.4 cases per 100 full-time workers]; retail trade = 0.68 [3.0]; manufacturing = 0.61 [2.7]; construction/extraction = 0.50 [2.2]; professional/scientific/technical services = 0.16 [0.7])[20]. Using occupational injury/illness incidence as a proxy for each sector's productivity sensitivity to averted community-level DALYs is a modeling simplification pending direct empirical validation; the underlying incidence rates are drawn from published national data, and the resulting weights are user-adjustable within the EJT interface, consistent with the treatment of other EJIP sub-component parameters.

**Integration into EJIP Priority Score:** The Health(u) component of the EJIP formula (Section 4.1) aggregates three normalized sub-components—DALY reduction, ROI, and time savings—into a single composite score. Sub-component weights are pre-specified in the study analysis plan and are user-adjustable within the EJT interface, allowing sensitivity analyses with alternative weighting schemes. Each sub-component is normalized to [0, 1] across all census tracts in the analysis region before aggregation, ensuring dimensional consistency within the EJIP weighted sum.

#### 4.2 Uncertainty, Bias, and Risk Quantification (UBR)

**Novelty of the UBR approach:** To our knowledge, the EJT UBR module represents the first implementation of real-time algorithmic bias monitoring with pre-specified acceptance criteria in an educational technology intervention delivered in K–12 classrooms. Existing educational AI tools (intelligent tutoring systems, automated essay scoring, recommendation engines) monitor accuracy and completion metrics but do not routinely monitor demographic parity of outputs during deployment. The UBR module fills this gap by computing demographic parity (DP) ratios across four protected groups (race/ethnicity, income quartile, linguistic isolation, disability status) at each EJT session, triggering a real-time alert when DP falls outside [0.8, 1.2] for any group. This threshold is grounded in the U.S. Equal Employment Opportunity Commission's 80% rule for adverse impact and the European Union AI Act's proposed non-discrimination standards for high-risk AI systems. All DP monitoring events, threshold crossings, and researcher responses are logged in the SHA-256 audit trail and reported in aggregate in the primary outcomes paper, providing a transparency standard that exceeds current practice in educational technology research.

**Purpose:** Propagate uncertainty through all calculations using Monte Carlo simulation, monitor for algorithmic bias using Demographic Parity (DP) metrics, and maintain complete audit trails for transparency and reproducibility.

**Monte Carlo Uncertainty Propagation (N = 10,000 iterations):**

1. Sample confidence scores: $c_i^{(k)} \sim \text{Beta}(\alpha_i, \beta_i)$ parameterized from point estimate
2. Sample dose-response parameters: $\beta^{(k)} \sim N(\hat{\beta}, SE^2)$ from literature uncertainty
3. Sample SVI measurement error: $SVI^{(k)} \sim N(SVI, 0.05^2)$
4. Compute $Priority^{(k)}(u)$ for all census tracts u using sampled parameters
5. Record rankings $R^{(k)} = rank(Priority^{(k)})$

**Output:** 95% credible intervals $[P_{2.5}, P_{97.5}]$ and rank stability = P(u in top-5 | u in top-5 at baseline)

**Demographic Parity Calculation:**

$$DP = P(HighPriority \mid ProtectedGroup) / P(HighPriority \mid ReferenceGroup)$$

**Acceptance criterion:** $DP \in [0.8, 1.2]$ for each protected demographic group
**Protected groups monitored:** Race/ethnicity, income quartile, linguistic isolation, disability status

### 5. 3+3 VALIDATION FRAMEWORK

To our knowledge, the 3+3 Validation Framework represents the first published validation structure for an AI-integrated environmental health decision support tool that explicitly separates output quality validation (are the EJT's investment priorities meaningful?) from system integrity verification (is the EJT operating as specified?). Existing AI validation frameworks in health contexts (e.g., TRIPOD-AI, PROBAST-AI) focus primarily on predictive model performance and do not address the multi-objective optimization, demographic parity monitoring, and audit trail completeness requirements of a cyber-physical decision support system. The 3+3 structure was designed specifically to address these gaps and is intended as a transferable framework for future AI-integrated health equity tools.

The EJT employs a 3+3 Validation Framework consisting of 3 primary validation metrics (testing whether outputs are meaningful) and 3 supporting system checks (testing whether the system operates correctly).

**Table 6. 3+3 Validation Framework Metrics**

| Metric | Threshold | Type | Rationale |
|---|---|---|---|
| **PRIMARY VALIDATION METRICS (Output Quality)** | | | |
| Expert Agreement (Spearman ρ) | **ρ > 0.60** | Primary | Large effect size (Cohen) |
| Rank Stability | **> 75%** | Primary | Majority stability |
| Theme Extraction Quality | **> 80%** | Primary | Exceeds human κ |
| **SUPPORTING SYSTEM CHECKS (Technical Function)** | | | |
| System Completion Rate | **≈ 100%** | Supporting | All inputs processed |
| Audit Trail Completeness | **= 100%** | Supporting | Full provenance |
| SVI Quartile Representation | **All quartiles** | Supporting | Equity coverage |

#### 5.1 Complete Validation Metrics (11 Total)

**Table 7. Complete Validation Metrics (11 Total)**

| # | Metric | Target | Module | Question Answered |
|---|---|---|---|---|
| 1 | CVET Score | Generated | CVET | Does LLM extract themes? |
| 2 | ESHI Score | Generated | ESHI | Do DRFs calculate benefits? |
| 3 | EJIP Priority Rank | Generated | EJIP | Does system produce rankings? |
| 4 | Bootstrap 95% CI | Computed | UBR | Are scores uncertain? |
| 5 | Rank Stability | **> 75%** | UBR | Are rankings robust? |
| 6 | Expert Agreement | **ρ > 0.60** | Validation | Do outputs match experts? |
| 7 | Audit Completeness | **100%** | UBR | Is there full traceability? |
| 8 | System Usability Scale (SUS) | **> 68** | Usability | Can users operate system? |
| **9** | **Theme Extraction Quality** | **> 80%** | CVET | Are LLM themes valid? |
| **10** | **Processing Time** | **< 5 min** | System | Is system efficient? |
| **11** | **Test-Retest Reliability** | **Intraclass Correlation Coefficient (ICC) > 0.80** | CVET | Are LLM outputs consistent? |

**Note:** *Green rows (9-11) are additions to strengthen validation. Metrics 1-8 are original.*

#### 5.2 Sample Size and Power Parameters

The following parameters justify the enrolled sample size required to achieve the validation thresholds specified in Section 5.1. **Figure 1** illustrates statistical power as a function of sample size and effect size for the individual-randomized design, providing the basis for the sample size calculation (n=26 per arm, 80% power, Hedges' g=0.80).

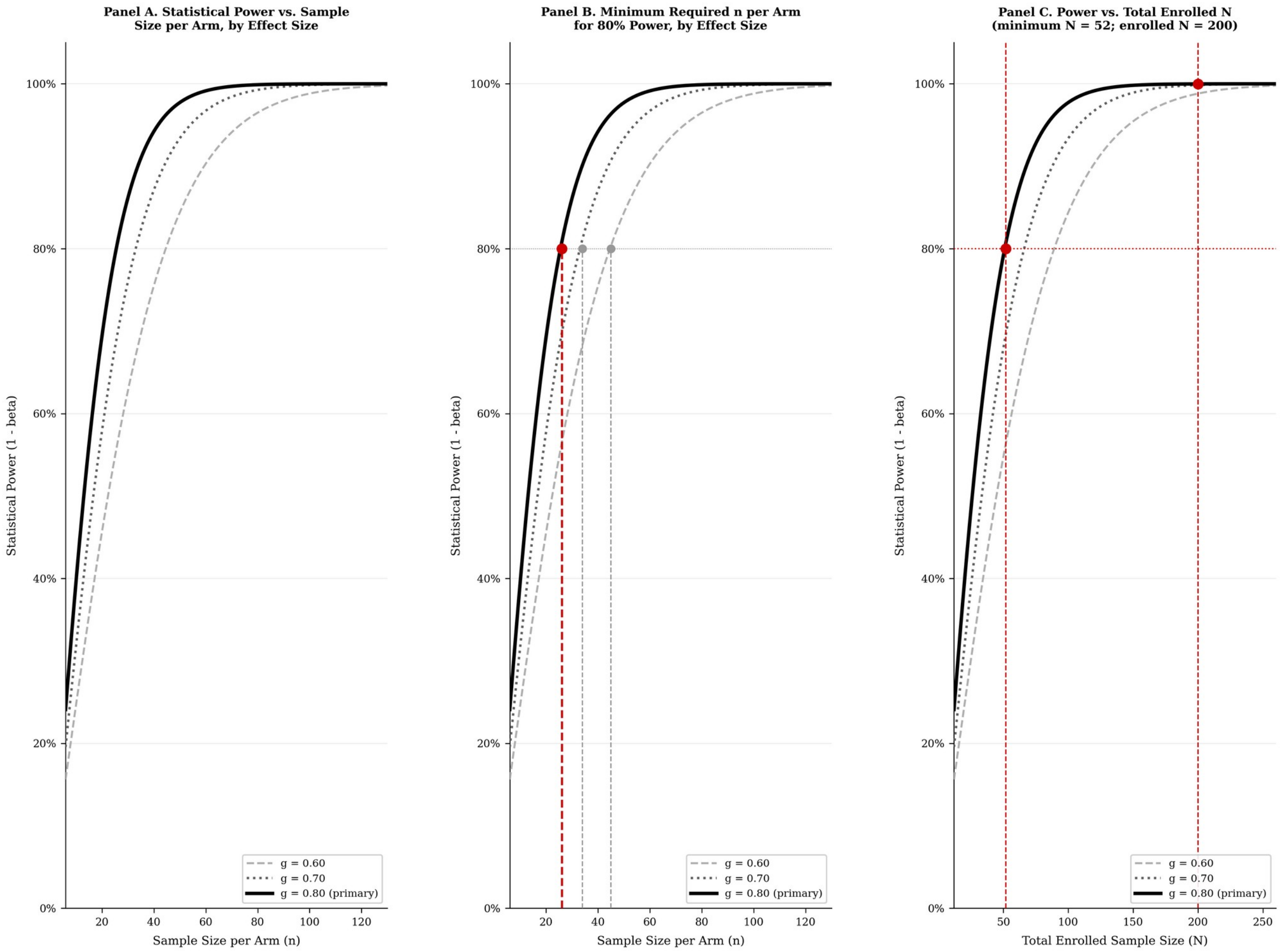


**Figure 1.** Statistical power and sample size parameters for the Ecosystem Justice Translator randomized controlled trial (Section 5.2).

**Panel A** shows statistical power as a function of sample size per arm for three effect sizes (Hedges' g = 0.60, 0.70, 0.80) under individual randomization (two-tailed α = 0.05, independent samples t-test, G*Power 3.1.9.7). **Panel B** shows the same power curves with the minimum required n per arm for 80% power marked at each effect size threshold. **Panel C** shows statistical power as a function of total enrolled sample size for three effect sizes (Hedges' g = 0.60, 0.70, 0.80) under two-tailed α = 0.05, illustrating the power gained by enrolling N = 200 beyond the minimum required n = 52.

Red lines and filled circles mark the study design assumptions: Minimum required: n = 26 per arm (G*Power 3.1.9.7; independent samples t-test; two-tailed α = 0.05; 80% power; Hedges' g = 0.80). Enrolled N = 200 (n = 100 per arm) provides >99% power at Hedges' g = 0.80; over-recruitment target N = 250 (25% buffer). Dashed line indicates minimum n per arm for 80% power at g = 0.80 (n = 26). Effect sizes reported as Hedges' g with J bias-correction (J ≈ 0.996 for N = 200, df = 198).[21] See **Section 5.1** for psychometric validation thresholds.

***— End of Technical Supplement —***
