## Supplementary material for "Integrating planetary health and environmental justice into high school construction career education: protocol for a randomized controlled trial of the Ecosystem Justice Translator": SPIRIT Checklist File

### Standard Protocol Items: Recommendations for Interventional Trials (SPIRIT) 2013 Checklist

Recommended Items to Address in a Clinical Trial Protocol

*Supplementary File 1*

#### COMPLIANCE SUMMARY: 33/33 items addressed (100%)

| Item | Description | Page | ✓ |
|---|---|---|---|
| **ADMINISTRATIVE INFORMATION** | | | |
| 1 | Title: Descriptive title identifying study design, population, interventions, and primary outcome | 1 | ✓ |
| 2a | Trial registration: Registry name and registration number | 2 (Abstract) | ✓ |
| 2b | All items from WHO Trial Registration Data Set | ClinicalTrials.gov | ✓ |
| 3 | Protocol version: Date and version identifier | 2 (Abstract) | ✓ |
| 4a | Funding: Sources of financial support | Funding (15) | ✓ |
| 4b | Funding: Role of funders in study design, conduct, analysis | Funding (15) | ✓ |
| 5a | Names, affiliations, roles of protocol contributors | Title page | ✓ |
| 5b | Name and contact for trial sponsor | Investigator-initiated | ✓ |
| 5c | Role of sponsor/funder in study design and dissemination | Funding (15) | ✓ |
| 5d | Trial contact information for public and scientific queries | Title page, Contacts | ✓ |
| **INTRODUCTION** | | | |
| 6a | Background: Description of research question with references to current knowledge | Introduction (2-3) | ✓ |
| 6b | Background: Explanation for choice of comparators | Introduction (5) | ✓ |
| 7 | Objectives: Specific objectives or hypotheses | Introduction (5) | ✓ |
| 8 | Trial design: Type of trial, allocation ratio, and framework | Methods (6) | ✓ |
| **METHODS: PARTICIPANTS, INTERVENTIONS, OUTCOMES** | | | |
| 9 | Study setting: Description of sites and relevant demographics | Methods (6) | ✓ |
| 10 | Eligibility criteria: Inclusion and exclusion criteria for participants | Methods (7) | ✓ |
| 11a | Interventions: Precise details of interventions for each group | Methods (7-8) | ✓ |
| 11b | Interventions: Criteria for discontinuing or modifying interventions | Methods (7), Ethics (14) | ✓ |
| 11c | Interventions: Strategies to improve adherence and monitoring | Methods (8) | ✓ |
| 11d | Interventions: Relevant concomitant care permitted or prohibited | Methods (8) | ✓ |
| 12 | Outcomes: Primary, secondary outcomes with timing of assessment | Methods (8-9) | ✓ |
| 13 | Participant timeline: Schedule of enrollment, interventions, assessments | Table 2 | ✓ |
| 14 | Sample size: Number to be enrolled and determination method | Methods (11) | ✓ |
| 15 | Recruitment: Strategies for achieving adequate enrollment | Methods (12) | ✓ |
| **METHODS: ASSIGNMENT OF INTERVENTIONS** | | | |
| 16a | Allocation: Sequence generation method | Methods (12) | ✓ |
| 16b | Allocation: Concealment mechanism | Methods (12) | ✓ |
| 16c | Allocation: Implementation (who generates sequence, enrolls, assigns) | Methods (12) | ✓ |
| 17a | Blinding: Who will be blinded and procedures for blinding | Methods (12) | ✓ |
| 17b | Blinding: Circumstances for unblinding if applicable | N/A (open-label) | ✓ |
| **METHODS: DATA COLLECTION, MANAGEMENT, ANALYSIS** | | | |
| 18a | Data collection: Plans for assessment and collection of outcome data | Methods (12-13) | ✓ |
| 18b | Data collection: Plans to promote participant retention | Methods (12) | ✓ |
| 19 | Data management: Plans for data entry, coding, security, storage | Methods (12-13) | ✓ |
| 20a | Statistical methods: Methods for primary and secondary outcomes | Methods (13) | ✓ |
| 20b | Statistical methods: Methods for additional analyses (subgroup, adjusted) | Methods (13) | ✓ |
| 20c | Statistical methods: Definition of analysis population and missing data | Methods (13) | ✓ |
| **METHODS: MONITORING** | | | |
| 21a | Data monitoring: Composition of DMC or explanation why not needed | Methods (13) | ✓ |
| 21b | Data monitoring: Interim analyses and stopping guidelines | Methods (13) | ✓ |
| 22 | Harms: Plans for collecting, assessing, reporting adverse events | Methods (13) | ✓ |
| 23 | Auditing: Frequency and procedures for trial auditing | Methods (12) | ✓ |
| **ETHICS AND DISSEMINATION** | | | |
| 24 | Research ethics approval: Plans for obtaining approval | Ethics (14) | ✓ |
| 25 | Protocol amendments: Plans for communicating amendments | Ethics (14) | ✓ |
| 26a | Consent: Who will obtain consent and how | Ethics (14) | ✓ |
| 26b | Consent: Additional consent provisions for collection of biological specimens | N/A | ✓ |
| 27 | Confidentiality: Data protection procedures | Ethics (14) | ✓ |
| 28 | Declaration of interests: Financial and other competing interests | Competing Interests (15) | ✓ |
| 29 | Access to data: Who will have access to final trial dataset | Data Availability (15) | ✓ |
| 30 | Post-trial care: Provisions for post-trial care (if applicable) | Ethics (14) | ✓ |
| 31a | Dissemination: Plans for investigator communication of trial results | Ethics (14) | ✓ |
| 31b | Dissemination: Authorship eligibility guidelines and use of writers | Ethics (14) | ✓ |
| 31c | Dissemination: Plans for granting public access to protocol, data | Ethics (14), Data | ✓ |

| Item | Description | Page | ✓ |
| --- | --- | --- | --- |
| | | Availability (15) | |
| **APPENDICES** | | | |
| 32 | Informed consent materials | Supplementary File 4 (Stanford IRB eProtocol #84369, approved February 13, 2026) | ✓ |
| 33 | Biological specimens (if applicable) | N/A | ✓ |

### SPIRIT-Artificial Intelligence (AI) 2020 Extension

Additional Items for AI Intervention Trials

*Reference: Rivera SC et al. Nat Med 2020;26:1351-63*

| Item | Description | Page | ✓ |
|---|---|---|---|
| **SPIRIT-AI EXTENSION ITEMS** | | | |
| AI-1 | State that the intervention uses AI/ML and specify the type of AI system | Methods (7) | ✓ |
| AI-2 | Describe AI system inputs, outputs, and clinical workflow integration | Methods (7-8), Supp 2 | ✓ |
| AI-3 | Describe how the AI system handles the data (preprocessing, analysis) | Supp File 2 | ✓ |
| AI-4 | Describe the AI system's training data and development process | Methods (7), Supp 2 | ✓ |
| AI-5 | Describe any human-AI interaction and decision-making process | Methods (8) | ✓ |
| AI-6 | Describe error analysis plans and handling of AI failures | Methods (7), Supp File 2 | ✓ |
| AI-7 | Describe version control and any planned updates during trial | Methods (7) | ✓ |
| AI-8 | Consider equity implications of the AI intervention | Methods (7-8), PPI (15) | ✓ |
| AI-9 | Describe plans for assessing algorithmic bias | Supp File 2, UBR | ✓ |

#### COMPLIANCE SUMMARY

This protocol addresses all 33 SPIRIT 2013 items and all 9 SPIRIT-AI extension items applicable to AI-based educational interventions.

##### Additional Guidelines Followed:

- CONSORT-Equity 2017 Extension for health equity reporting
- TIDieR Checklist for intervention description
- ICMJE authorship criteria

*--- End of Checklist ---*
